# Clinical predictors of COVID-19 mortality

**DOI:** 10.1101/2020.05.19.20103036

**Authors:** Arjun S. Yadaw, Yan-chak Li, Sonali Bose, Ravi Iyengar, Supinda Bunyavanich, Gaurav Pandey

## Abstract

**Background:** The coronavirus disease 2019 (COVID-19) pandemic has affected over millions of individuals and caused hundreds of thousands of deaths worldwide.

It can be difficult to accurately predict mortality among COVID-19 patients presenting with a spectrum of complications, hindering the prognostication and management of the disease.

**Methods:** We applied machine learning techniques to clinical data from a large cohort of 5,051 COVID-19 patients treated at the Mount Sinai Health System in New

York City, the global COVID-19 epicenter, to predict mortality. Predictors were designed to classify patients into Deceased or Alive mortality classes and were evaluated in terms of the area under the receiver operating characteristic (ROC) curve (AUC score).

**Findings:** Using a development cohort (n=3,841) and a systematic machine learning framework, we identified a COVID-19 mortality predictor that demonstrated high accuracy (AUC=0·91) when applied to test sets of retrospective (n= 961) and prospective (n=249) patients. This mortality predictor was based on five clinical features: age, minimum O_2_ saturation during encounter, type of patient encounter (inpatient vs. various types of outpatient and telehealth encounters), hydroxychloroquine use, and maximum body temperature.

**Interpretation:** An accurate and parsimonious COVID-19 mortality predictor based on five features may have utility in clinical settings to guide the management and prognostication of patients affected by this disease.

**Funding:** This work was funded by the National Institutes of Health.

## Introduction

The coronavirus disease 2019 (COVID-19) pandemic has affected over 3.6 million individuals, and caused over 250,000 deaths worldwide as of May 5^th^, 2020.(1) Although the causative SARS-CoV-2 virus primarily targets the respiratory system(2, 3), complications in other organ systems, e.g., cardiovascular, neurologic and renal, can also contribute to death from the disease. Clinical experience thus far has demonstrated significant heterogeneity in the trajectory of SARS-CoV-2 infection, spanning patients who are asymptomatic to those with mild, moderate, and severe disease forms, with a high percentage of patients who do not survive(2, 3). Notably, it can be difficult to accurately predict clinical outcomes for patients across this spectrum of clinical presentations. This presents an enormous challenge to the prognostication and management of COVID-19 patients, especially within disease epicenters such as New York City (NYC) that need to triage a high volume of patients. Accurate prediction of COVID-19 mortality, and the identification of contributing factors would therefore allow for targeted strategies in patients with the highest risk of death.

Towards this aim, we analyzed clinical data from 5051 patients who had laboratory confirmed COVID-19 and were treated within multiple hospitals and locations of the Mount Sinai Health System spanning different boroughs of NYC. We used multiple machine learning-based classification algorithms(4) to develop models that can accurately predict mortality from COVID-19. We also identified clinical features that contributed the most to this prediction. An improved understanding of predictive factors for COVID-19 is critical to the development of clinical decision support systems that can better identify those with higher risk of mortality, and inform interventions to reduce the risk of death.

## Methods

### Study population

De-identified electronic medical record (EMR) data from patients diagnosed with COVID-19 within the Mount Sinai Hospital System, New York, NY through April 7, 2020 were included in the study. The Mount Sinai Health System is a network of 5 hospital campuses and over 400 ambulatory practices spanning the New York metropolitan area (Supplementary Table 1). COVID-19 diagnosis was based on positive polymerase chain reaction (PCR)-based clinical laboratory testing for the SARS-CoV-2 virus.

Data from COVID-19 patients through April 6, 2020 were randomly split into two groups of independent subjects comprising 80% of the sample (n=3841) for development of the mortality predictor (i.e. development set), and 20% (n=961) to serve as retrospective test set 1. A prospective validation set of independent subjects, test set 2, included COVID-19 patients encountered on April 7, 2020 (n=249).

### Identification and validation of the predictor

We implemented a systematic machine learning-based framework to construct the mortality predictor from the development set using missing value imputation(5), feature selection(6), classification(4) and statistical(7) techniques. The goal of this predictor was to classify a COVID-19 patient as likely to survive or die from the disease, i.e., “Alive” or “Deceased,” respectively. The identified predictor was then validated in test sets 1 and 2 in terms of the Area Under the ROC Curve (AUC score)(8). The overall workflow is shown in Figure 1, and detailed methods are provided in Supplementary Material.

**Figure 1:**
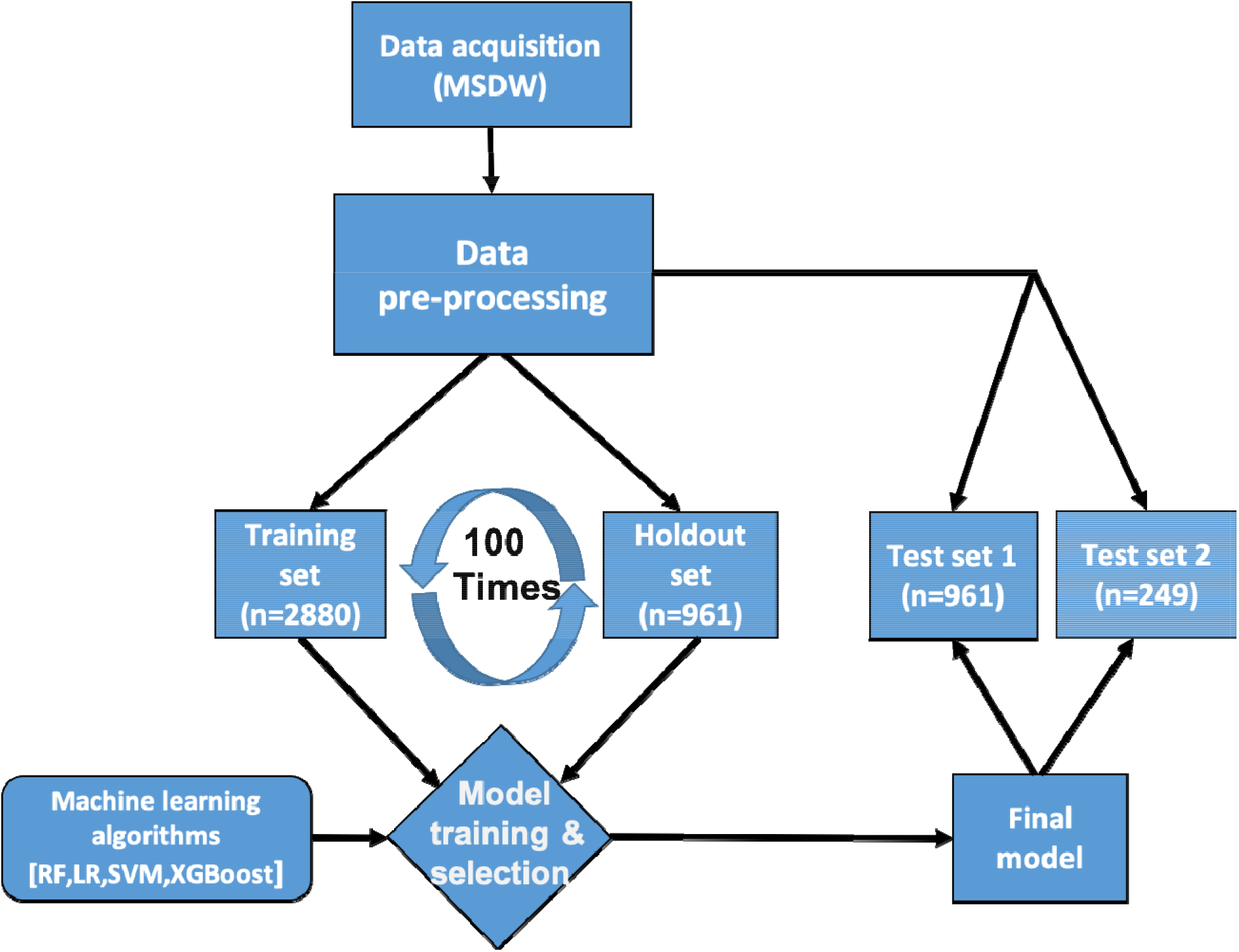
Workflow for data management and COVID-19 mortality predictor development. Data were obtained from the Mount Sinai Data Warehouse (MSDW). After pre-processing, COVID-19-positive patients’ data (n=4802) were randomly divided in an 80:20 ratio into a predictor development (n=3841) and an independent retrospective validation dataset (test set 1; n=961). For predictor training and selection, the development set was further randomly split into a 60% training dataset (n=2880) and a 20% holdout dataset (n=961). Four classification algorithms (logistic regression (LR), random forest (RF), support vector machine(SVM) and eXtreme Gradient Boosting (XGBoost)) were evaluated. The final predictive model was validated on test set 1 and another independent prospective validation set (test set 2; n=249). The complete details of the computational methods used can be found in Supplementary Material.

### Role of the funding source

The funding organizations had no role in the design and conduct of the study; collection, management, analysis, and interpretation of the data; preparation, review, or approval of the manuscript; and decision to submit the manuscript for publication.

## Results

### Patient characteristics

The demographic and clinical characteristics of the COVID-19 patients included in the development set (n=3841), test set 1 (n=961) and test set 2 (n=249) are shown in Table 1. The majority (55·3%) of patients in the development set were male, with an even higher prevalence of male sex among the deceased (61·3%). COVID-19 patients were mostly Caucasian (25·3%), African American (26·2%) and Latino(24·3%), with a minority identifying as Asian (4·2%). Hypertension and diabetes were the most common comorbidities (22·6% and 15·8%%, respectively). While a small minority were obese (6·0%) or had cancer (5·4%), an even smaller percentage had asthma (4·2%), COPD (2·3%), or currently smoked (3·5%). Over a third of the patients had been treated with azithromycin and/or hydroxychloroquine, consistent with the health system’s treatment practices during this time period.

**Table 1:**
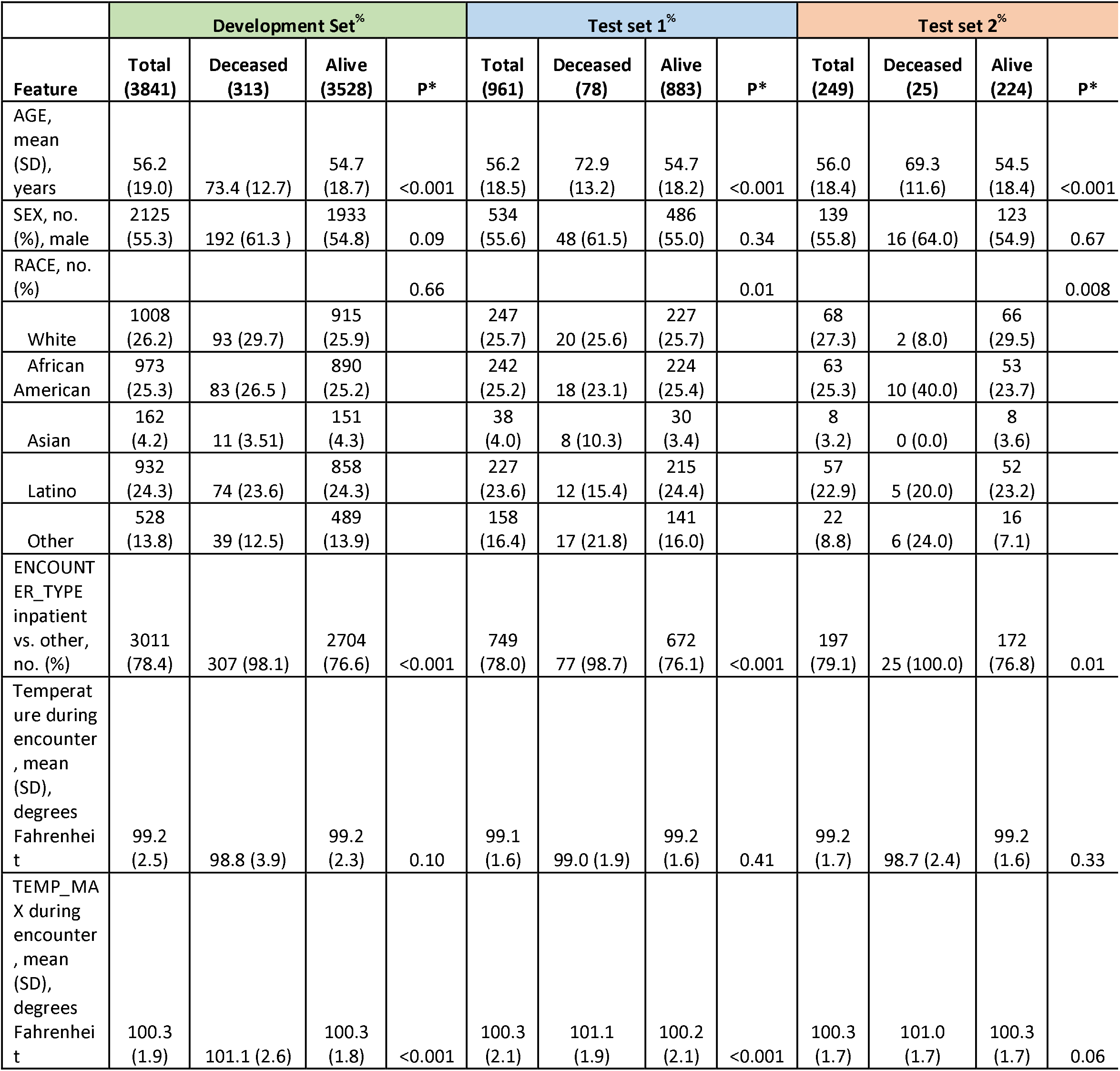
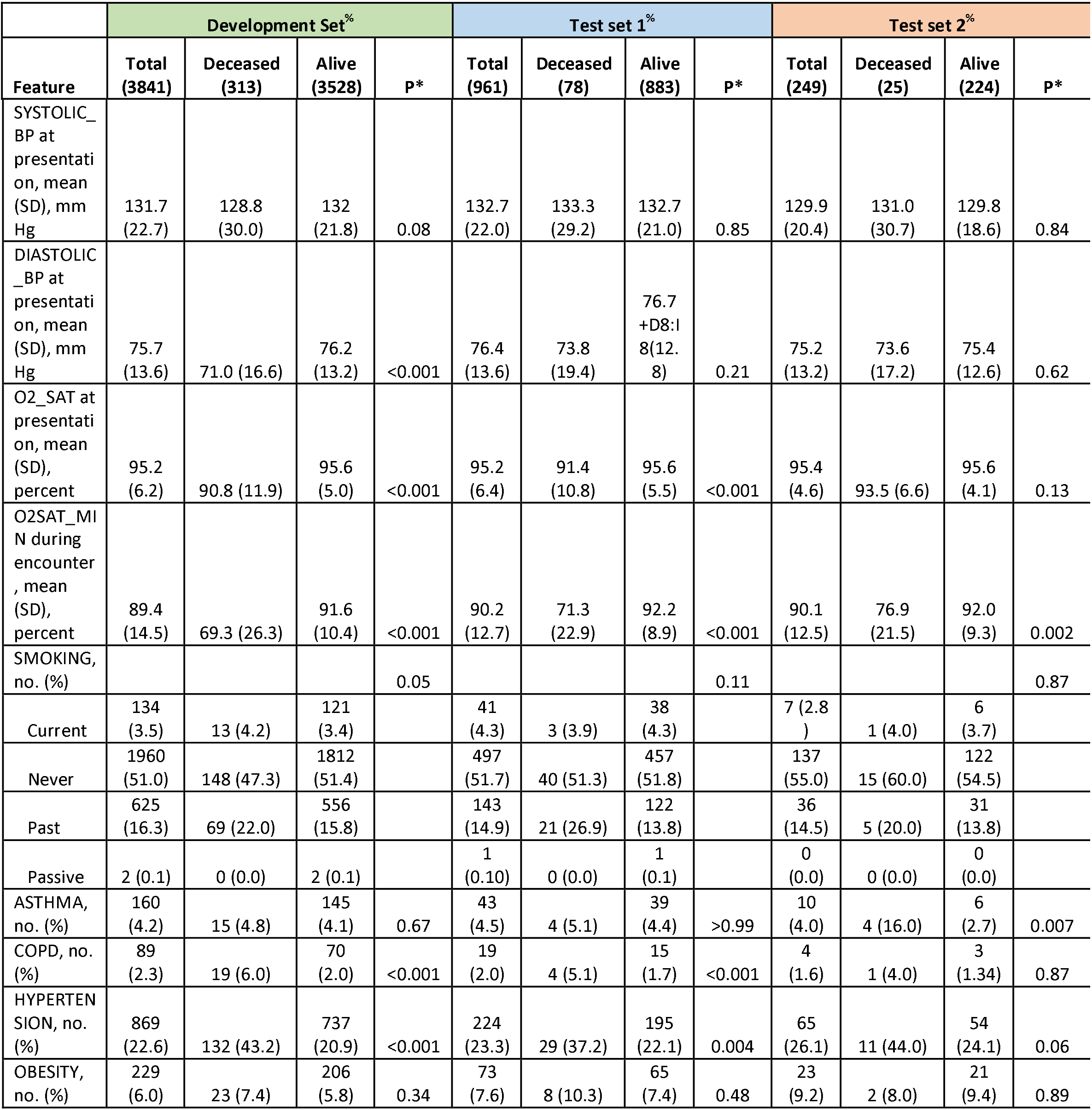
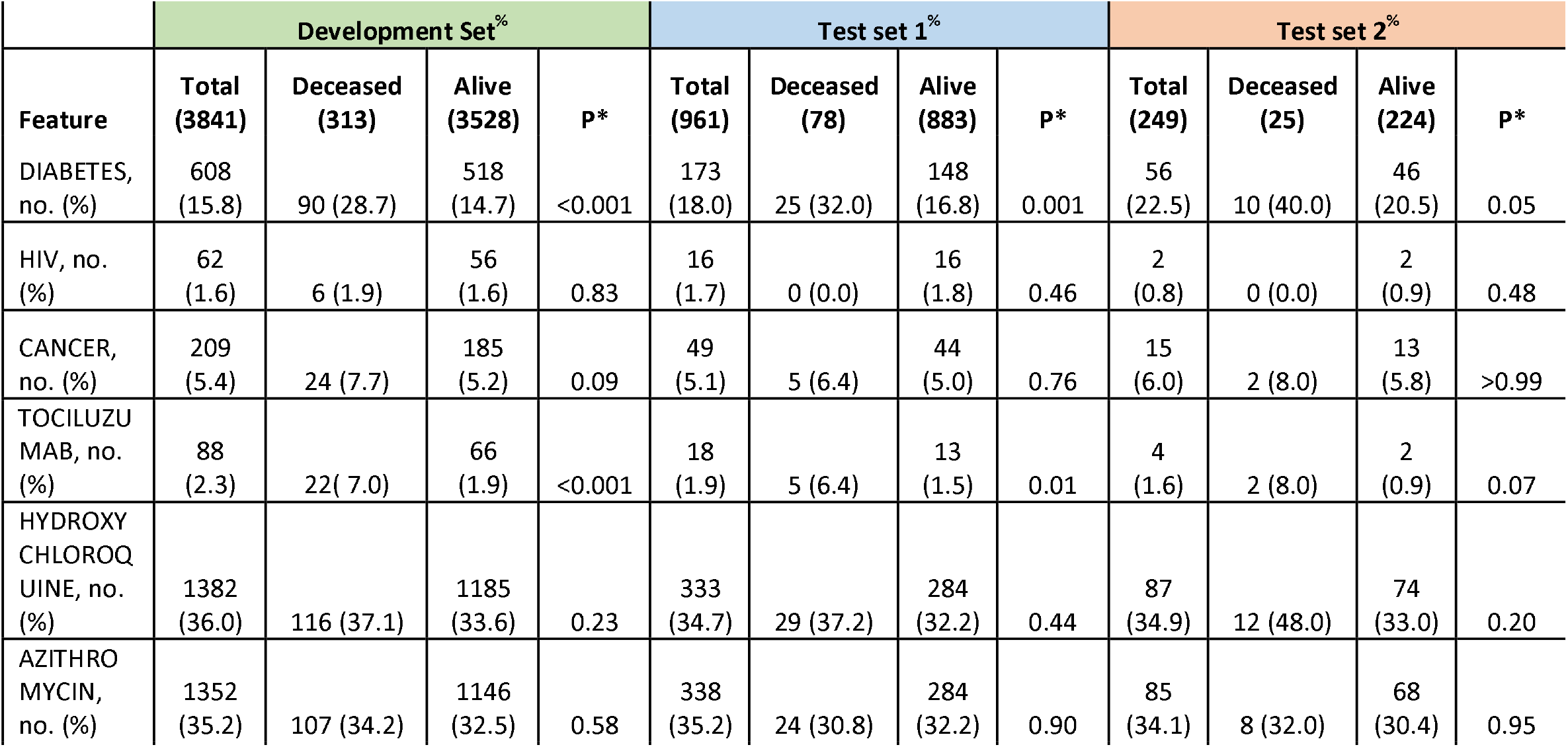
Characteristics of patients in the development and test sets. %Number of patients in each class in the corresponding set is shown in parentheses below the name of the class. *P(-values) from student’s t-test for continuous features and chi-square test for categorical features.

|  | Development Set<sup>%</sup> |  |  |  | Test set 1<sup>%</sup> |  |  |  | Test set 2<sup>%</sup> |  |  |  |
| --- | --- | --- | --- | --- | --- | --- | --- | --- | --- | --- | --- | --- |
| Feature | Total<br>(3841) | Deceased<br>(313) | Alive<br>(3528) | P* | Total<br>(961) | Deceased<br>(78) | Alive<br>(883) | P* | Total<br>(249) | Deceased<br>(25) | Alive<br>(224) | P* |
| AGE, mean (SD), years | 56.2 (19.0) | 73.4 (12.7) | 54.7 (18.7) | <0.001 | 56.2 (18.5) | 72.9 (13.2) | 54.7 (18.2) | <0.001 | 56.0 (18.4) | 69.3 (11.6) | 54.5 (18.4) | <0.001 |
| SEX, no. (%), male | 2125 (55.3) | 192 (61.3 ) | 1933 (54.8) | 0.09 | 534 (55.6) | 48 (61.5) | 486 (55.0) | 0.34 | 139 (55.8) | 16 (64.0) | 123 (54.9) | 0.67 |
| RACE, no. (%) |  |  |  | 0.66 |  |  |  | 0.01 |  |  |  | 0.008 |
| White | 1008 (26.2) | 93 (29.7) | 915 (25.9) |  | 247 (25.7) | 20 (25.6) | 227 (25.7) |  | 68 (27.3) | 2 (8.0) | 66 (29.5) |  |
| African American | 973 (25.3) | 83 (26.5 ) | 890 (25.2) |  | 242 (25.2) | 18 (23.1) | 224 (25.4) |  | 63 (25.3) | 10 (40.0) | 53 (23.7) |  |
| Asian | 162 (4.2) | 11 (3.51) | 151 (4.3) |  | 38 (4.0) | 8 (10.3) | 30 (3.4) |  | 8 (3.2) | 0 (0.0) | 8 (3.6) |  |
| Latino | 932 (24.3) | 74 (23.6) | 858 (24.3) |  | 227 (23.6) | 12 (15.4) | 215 (24.4) |  | 57 (22.9) | 5 (20.0) | 52 (23.2) |  |
| Other | 528 (13.8) | 39 (12.5) | 489 (13.9) |  | 158 (16.4) | 17 (21.8) | 141 (16.0) |  | 22 (8.8) | 6 (24.0) | 16 (7.1) |  |
| ENCOUNTER_TYPE inpatient vs. other, no. (%) | 3011 (78.4) | 307 (98.1) | 2704 (76.6) | <0.001 | 749 (78.0) | 77 (98.7) | 672 (76.1) | <0.001 | 197 (79.1) | 25 (100.0) | 172 (76.8) | 0.01 |
| Temperature during encounter , mean (SD), degrees Fahrenheit | 99.2 (2.5) | 98.8 (3.9) | 99.2 (2.3) | 0.10 | 99.1 (1.6) | 99.0 (1.9) | 99.2 (1.6) | 0.41 | 99.2 (1.7) | 98.7 (2.4) | 99.2 (1.6) | 0.33 |
| TEMP_MAX during encounter , mean (SD), degrees Fahrenheit | 100.3 (1.9) | 101.1 (2.6) | 100.3 (1.8) | <0.001 | 100.3 (2.1) | 101.1 (1.9) | 100.2 (2.1) | <0.001 | 100.3 (1.7) | 101.0 (1.7) | 100.3 (1.7) | 0.06 |
| Feature | Total<br>(3841) | Deceased<br>(313) | Alive<br>(3528) | P* | Total<br>(961) | Deceased<br>(78) | Alive<br>(883) | P* | Total<br>(249) | Deceased<br>(25) | Alive<br>(224) | P* |
| SYSTOLIC_<br>BP at<br>presentati<br>on, mean<br>(SD), mm<br>Hg | 131.7<br>(22.7) | 128.8<br>(30.0) | 132<br>(21.8) | 0.08 | 132.7<br>(22.0) | 133.3<br>(29.2) | 132.7<br>(21.0) | 0.85 | 129.9<br>(20.4) | 131.0<br>(30.7) | 129.8<br>(18.6) | 0.84 |
| DIASTOLIC_<br>BP at<br>presentati<br>on, mean<br>(SD), mm<br>Hg | 75.7<br>(13.6) | 71.0 (16.6) | 76.2<br>(13.2) | <0.001 | 76.4<br>(13.6) | 73.8<br>(19.4) | 76.7<br>+D8:l<br>8(12.<br>8) | 0.21 | 75.2<br>(13.2) | 73.6<br>(17.2) | 75.4<br>(12.6) | 0.62 |
| O2_SAT at<br>presentati<br>on, mean<br>(SD),<br>percent | 95.2<br>(6.2) | 90.8 (11.9) | 95.6<br>(5.0) | <0.001 | 95.2<br>(6.4) | 91.4<br>(10.8) | 95.6<br>(5.5) | <0.001 | 95.4<br>(4.6) | 93.5 (6.6) | 95.6<br>(4.1) | 0.13 |
| O2SAT_MI<br>N during<br>encounter<br>, mean<br>(SD),<br>percent | 89.4<br>(14.5) | 69.3 (26.3) | 91.6<br>(10.4) | <0.001 | 90.2<br>(12.7) | 71.3<br>(22.9) | 92.2<br>(8.9) | <0.001 | 90.1<br>(12.5) | 76.9<br>(21.5) | 92.0<br>(9.3) | 0.002 |
| SMOKING,<br>no. (%) |  |  |  | 0.05 |  |  |  | 0.11 |  |  |  | 0.87 |
| Current | 134<br>(3.5) | 13 (4.2) | 121<br>(3.4) |  | 41<br>(4.3) | 3 (3.9) | 38<br>(4.3) |  | 7 (2.8<br>) | 1 (4.0) | 6<br>(3.7) |  |
| Never | 1960<br>(51.0) | 148 (47.3) | 1812<br>(51.4) |  | 497<br>(51.7) | 40 (51.3) | 457<br>(51.8) |  | 137<br>(55.0) | 15 (60.0) | 122<br>(54.5) |  |
| Past | 625<br>(16.3) | 69 (22.0) | 556<br>(15.8) |  | 143<br>(14.9) | 21 (26.9) | 122<br>(13.8) |  | 36<br>(14.5) | 5 (20.0) | 31<br>(13.8) |  |
| Passive | 2 (0.1) | 0 (0.0) | 2 (0.1) |  | 1<br>(0.10) | 0 (0.0) | 1<br>(0.1) |  | 0<br>(0.0) | 0 (0.0) | 0<br>(0.0) |  |
| ASTHMA,<br>no. (%) | 160<br>(4.2) | 15 (4.8) | 145<br>(4.1) | 0.67 | 43<br>(4.5) | 4 (5.1) | 39<br>(4.4) | >0.99 | 10<br>(4.0) | 4 (16.0) | 6<br>(2.7) | 0.007 |
| COPD, no.<br>(%) | 89<br>(2.3) | 19 (6.0) | 70<br>(2.0) | <0.001 | 19<br>(2.0) | 4 (5.1) | 15<br>(1.7) | <0.001 | 4<br>(1.6) | 1 (4.0) | 3<br>(1.34) | 0.87 |
| HYPERTEN<br>SION, no.<br>(%) | 869<br>(22.6) | 132 (43.2) | 737<br>(20.9) | <0.001 | 224<br>(23.3) | 29 (37.2) | 195<br>(22.1) | 0.004 | 65<br>(26.1) | 11 (44.0) | 54<br>(24.1) | 0.06 |
| OBESITY,<br>no. (%) | 229<br>(6.0) | 23 (7.4) | 206<br>(5.8) | 0.34 | 73<br>(7.6) | 8 (10.3) | 65<br>(7.4) | 0.48 | 23<br>(9.2) | 2 (8.0) | 21<br>(9.4) | 0.89 |
| Feature | Total<br>(3841) | Deceased<br>(313) | Alive<br>(3528) | P* | Total<br>(961) | Deceased<br>(78) | Alive<br>(883) | P* | Total<br>(249) | Deceased<br>(25) | Alive<br>(224) | P* |
| DIABETES,<br>no. (%) | 608<br>(15.8) | 90 (28.7) | 518<br>(14.7) | <0.001 | 173<br>(18.0) | 25 (32.0) | 148<br>(16.8) | 0.001 | 56<br>(22.5) | 10 (40.0) | 46<br>(20.5) | 0.05 |
| HIV, no.<br>(%) | 62<br>(1.6) | 6 (1.9) | 56<br>(1.6) | 0.83 | 16<br>(1.7) | 0 (0.0) | 16<br>(1.8) | 0.46 | 2<br>(0.8) | 0 (0.0) | 2<br>(0.9) | 0.48 |
| CANCER,<br>no. (%) | 209<br>(5.4) | 24 (7.7) | 185<br>(5.2) | 0.09 | 49<br>(5.1) | 5 (6.4) | 44<br>(5.0) | 0.76 | 15<br>(6.0) | 2 (8.0) | 13<br>(5.8) | >0.99 |
| TOCILUZU<br>MAB, no.<br>(%) | 88<br>(2.3) | 22 (7.0) | 66<br>(1.9) | <0.001 | 18<br>(1.9) | 5 (6.4) | 13<br>(1.5) | 0.01 | 4<br>(1.6) | 2 (8.0) | 2<br>(0.9) | 0.07 |
| HYDROXY<br>CHLOROQ<br>UINE, no.<br>(%) | 1382<br>(36.0) | 116 (37.1) | 1185<br>(33.6) | 0.23 | 333<br>(34.7) | 29 (37.2) | 284<br>(32.2) | 0.44 | 87<br>(34.9) | 12 (48.0) | 74<br>(33.0) | 0.20 |
| AZITHRO<br>MYCIN,<br>no. (%) | 1352<br>(35.2) | 107 (34.2) | 1146<br>(32.5) | 0.58 | 338<br>(35.2) | 24 (30.8) | 284<br>(32.2) | 0.90 | 85<br>(34.1) | 8 (32.0) | 68<br>(30.4) | 0.95 |

Univariate analyses of patient characteristics in the development set (Table 1) showed that COVID-19 patients who died were significantly older with a mean age of 73·4 (SD 12·7) vs. 54·7 (SD 18·7) years in survivors (P<0·001). They were more likely to have had their initial encounter at a hospital rather than at an outpatient or telehealth setting within our hospital system (P<0·001). Those who died had higher body temperature and lower oxygen saturation at initial presentation, and their minimum oxygen (O_2_) saturation over the duration of their encounter was also lower (P<0·001 for all). Death from COVID-19 was associated with smoking (P=0·05), COPD (P<0·001), hypertension (P<0·001), and diabetes (P<0·001).

The characteristics of test sets 1 and 2 were largely similar to those of the development set, except for some differences in the relative proportions of race(Table 1). While minimum O_2_ saturation during encounter was consistently lower in the deceased vs. alive patients in both test sets, O_2_ saturation at presentation was lower among the deceased in test set 1 only. COPD, hypertension, and diabetes were more prevalent among the deceased in test set 1, but there were no significant differences in these comorbidities in test set 2.

### Development and validation of the predictor

Following imputation, there were twenty distinct clinical features with less than 20% missing values in the development set that improved predictor performance (Figure 2A). Compared to the other classification algorithms (LR, RF, SVM), XGBoost performed significantly better at this and higher levels of missing values(Figure 2A; Friedman-Nemenyi P<0·001). Therefore, we used the imputed version of the development set with 20 features and XGBoost, to develop the first COVID-19 mortality predictor in this study, referred to as the 20F model.

**Figure 2:**
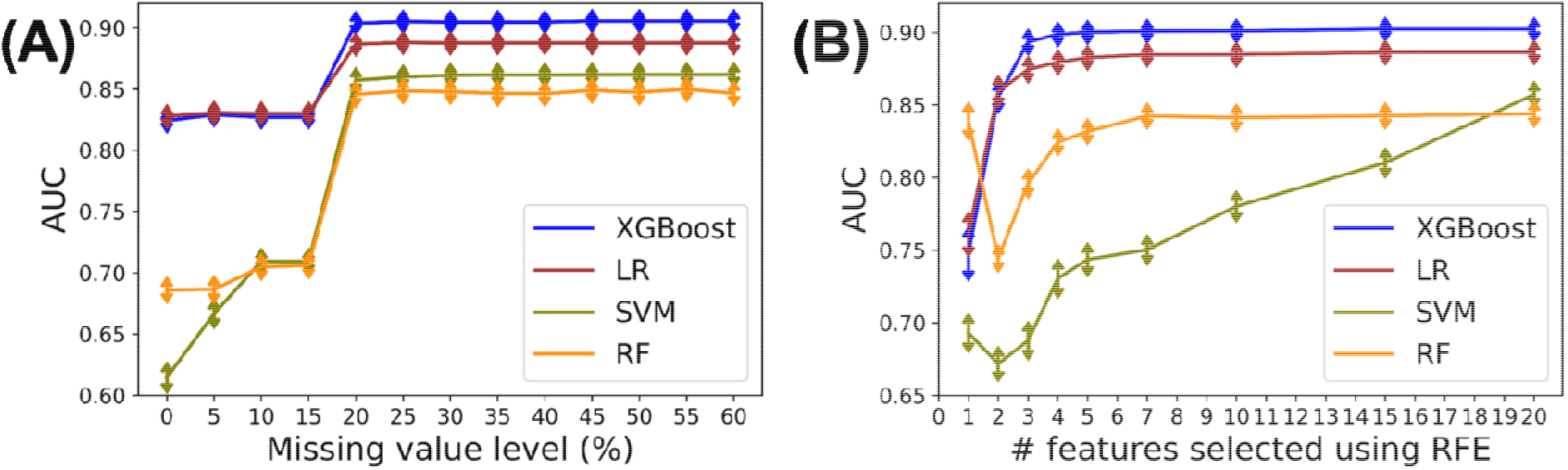
Results from missing value imputation and feature selection during predictor training and selection. **(A)** We attempted to find the optimal level of missing values in the range of 0% to 60% that could be imputed and lead to more accurate predictors. For this, we took incremental steps of 5% in missing value levels, and used mean and mode imputation for continuous and categorical features respectively. At each level, four candidate classifiers (LR, RF, SVM and XGBoost) were trained and evaluated on the corresponding holdout set in terms of the area under the receiver operating characteristic (ROC) curve (AUC score) as the metric. This process was repeated 100 times and the average AUCs for each candidate classifier and missing value level are shown here, along with error bars denoted by vertical arrows. (B) Using a setup analogous to (A), and the Recursive Feature Elimination(RFE) algorithm, we evaluated the performance of the four classification algorithms with different number of features selected from the full set of twenty. The average AUC scores from 100 runs of this process are shown here, along with error bars denoted by vertical arrows. The details of the computational methods underlying these analyses are provided in Supplementary Material. LR=logistic regression; RF=random forest; SVM=support vector machine.

We also tested if a smaller subset of the 20 features could yield an even more accurate predictor, since such a subset would be easier to study and implement in a clinical setting. Indeed, we found that for the best-performing XGBoost algorithm(Friedman-Nemenyi P<0·001), the AUC saturated at as few as five features (Figure 2B), validating our hypothesis that fewer than 20 features could yield an accurate predictor. The five features identified from the development set included the following: minimum oxygen saturation recorded during the encounter, patient’s age, type of encounter, maximum body temperature during the encounter, and use of hydroxychloroquine during treatment. We trained this second Covid-19 mortality predictor, referred to as the 5F model, by applying XGBoost to these 5 features in the imputed development set.

Validation of the 20F and 5F models on test set 1 (retrospective data) and test set 2 (prospective data) both yielded very good performance (AUC>0·9; Figure 3). The predictor’s strong performance in both test sets demonstrated that the predictors constructed from data on a given day can be reliably applied retrospectively and prospectively.

**Figure 3:**
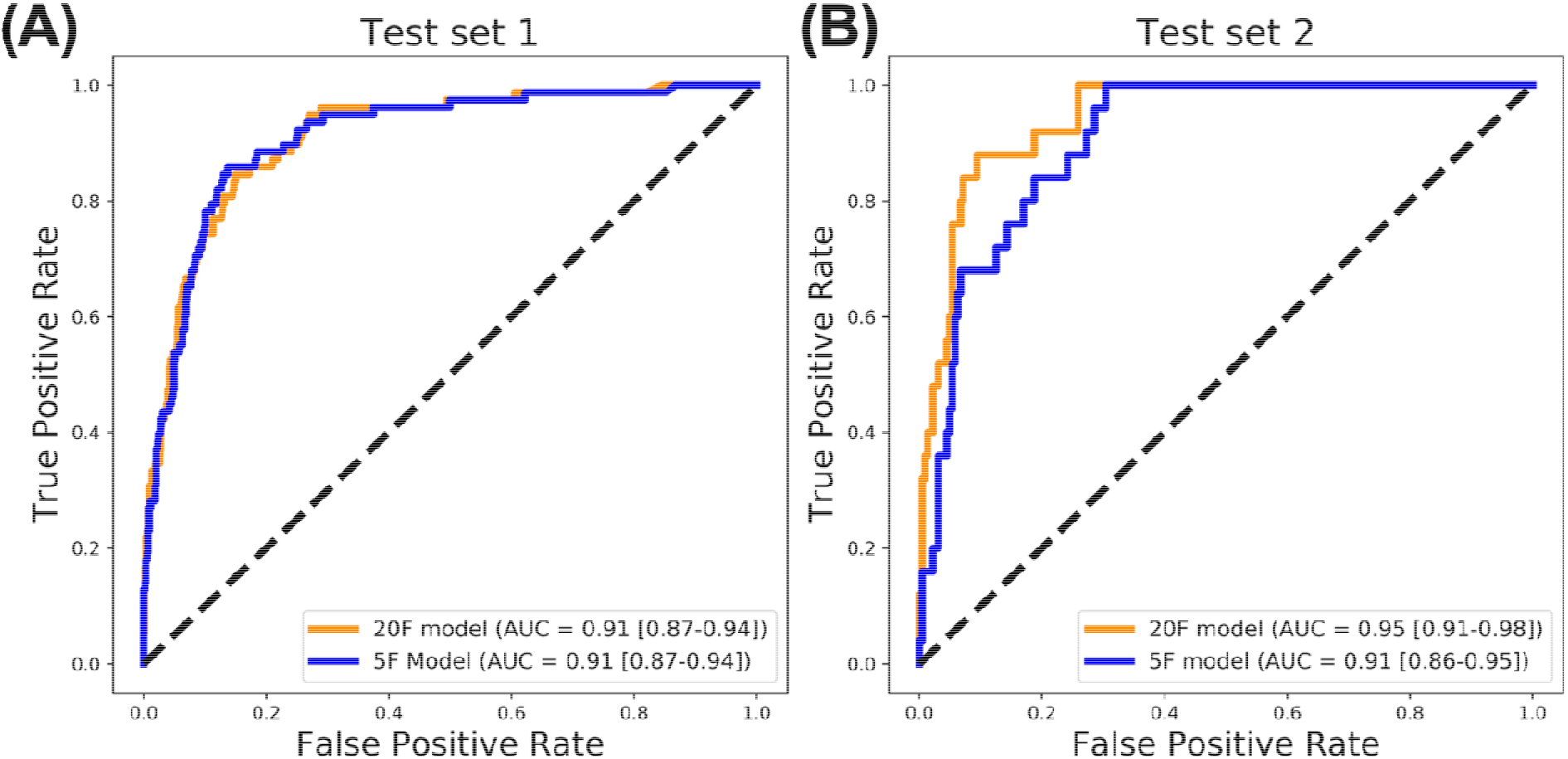
Performance of the final mortality predictors on two validation sets. Based on the results in Figure 2, we constructed two predictors: (1) Training XGBoost, the best performer in Figure 2(A), on 20 features with at most 20% missing values (20F model), and (2) Training XGBoost, the best performer in Figure 2(B), on the optimal 5 features at which the performance saturated (5F model). Both these predictors were evaluated on the (A) retrospective Test set 1 (n=961) and the(B) prospective Test set 2 (n=249). Evaluation results are shown here in terms of the ROC curves obtained, as well as their area under the curve (AUC) scores. The 95% confidence intervals of the AUC scores are shown in square brackets.

### Features predictive of mortality

Similar to the features that the 5F model was based on, we identified the five most predictive features for the other classification algorithms we tested (Figure 4A). While there was variability among these features due to the inherent differences among the algorithms, the age of the patient, and their minimum oxygen saturation level during the clinical encounter (O_2_SAT_min) were consistent across the algorithms. The values of O_2_SAT_min and age were indeed significantly different between the Deceased and Alive classes (Table 1, Figures 4B and 4C respectively; T-test P<0.001 for both features), affirming their predictive power. Supplementary Figure 1 shows that the top five features are consistent across all three runs of the feature selection and predictor development process.

**Figure 4:**
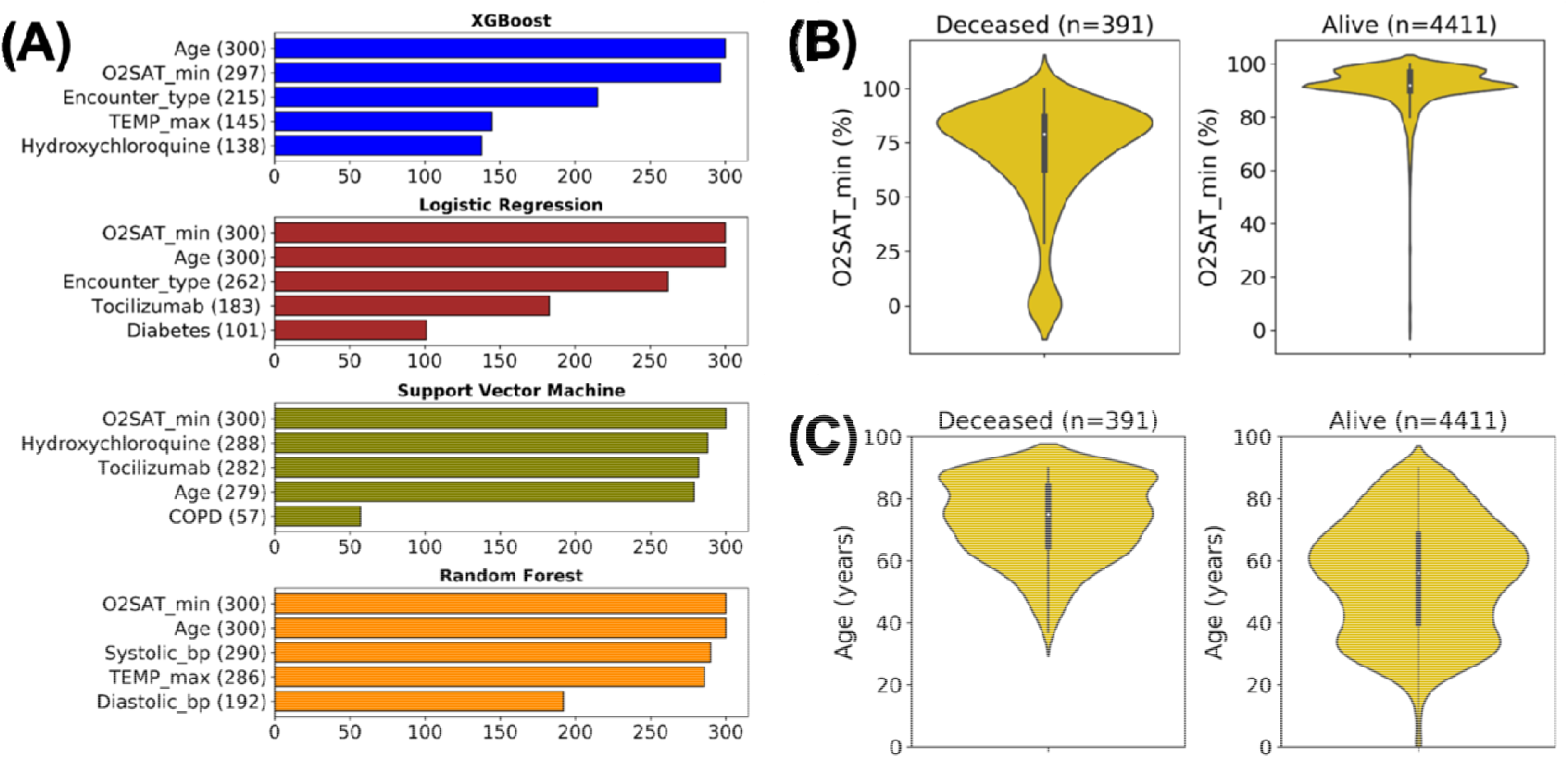
Top predictive features selected for the four classification algorithms tested. **(A)** Top five predictive features identified using the RFE algorithm for the four classification algorithms across three independent sets of hundred runs each of the predictor training and selection process described in Figures 1 and 2, and Supplementary Material. The values in parentheses indicate the number of times the feature was selected as top ranked. Also shown are violin plots representing the distributions of the values of the (B) O2SAT_min and (C) age features that were selected as top predictive features for all the four algorithms. The plot in (B) shows that the median value of O2SAT_min for the deceased group (79) was significantly lower (T-test 0.001) than that for the live group (92). Similarly, the plot in (C) shows that the median age in the deceased group (75) is significantly higher (T-test 0.001) than that in the alive group (56).

## Discussion

In this study, machine learning algorithms were applied to clinical and demographic data from 3841 COVID-19 patients from a major New York metropolitan area health system to identify and test a mortality predictor that demonstrated high accuracy(AUC=0·91) when applied to test sets of retrospective (n=961) and prospective(n=249) patient data. This mortality predictor was based on five clinical features: age, minimum O_2_ saturation, type of patient encounter (inpatient vs. outpatient and telehealth encounters), hydroxychloroquine use, and maximum body temperature. Given the heterogeneity in clinical presentation and course observed among COVID-19 patients,(2, 3) factors that contribute most to mortality are not always readily apparent, rendering care and management of COVID-19 patients difficult in settings of finite health care resources. Our data-driven findings may help clinicians better recognize and prioritize the care of patients at greatest risk of death.

A major strength of this study is that it was based on recent data from thousands of COVID-19 patients encountered within a global disease epicenter (NYC), resulting in findings that are highly relevant to the current pandemic. The results are based on rigorous machine learning analyses powered by a robust sample of patients with laboratory-confirmed COVID and demonstrate the potential of these methods to identify factors predicting mortality within clinical settings. Application of machine learning enabled the identification of predictors based on the XGBoost algorithm(9), where the clinical features contributed to mortality in a non-linear fashion. These predictors performed with high accuracy (AUC=0·91–0·95) in two independent validation sets of COVID-19 patients. Furthermore, the 5F model based on only the five features listed above performed almost as well as the 20F model based on all the features. This indicates that accurate mortality predictions can be obtained from a more parsimonious model, facilitating more efficient implementation in clinical environments.

Age and minimum oxygen saturation during encounter (O_2_SAT_min) were the most predictive features not only for the XGBoost algorithm, but for all four mortality classifiers tested (Figure 4), emphasizing these features’ epidemiological and clinical relevance. Since the beginnings of this pandemic, older age has been recognized as a risk factor(10, 11). In New York State, patients 60 years and over represent nearly 85% of all deaths due to COVID-19(12), and similarly high rates of mortality among those of advanced age have been noted in other COVID-19 hotspots across the United States(13). In addition, the fundamental clinical presentation of COVID-19 patients across the pandemic has been respiratory symptoms associated with hypoxia, often leading to subsequent respiratory failure and requiring ventilator support and/or extracorporeal membrane oxygenation(14). This study’s finding that a patient’s minimum oxygen saturation (O_2_SAT_min) value during hospitalization was the strongest predictive feature of mortality (Figure 4) is in line with global epidemiologic observations that respiratory failure is the most common feature of critical illness and death in COVID-10 patients(15, 16).

In addition to age and oxygen saturation, other features in the mortality predictor are also consistent with clinical observations accumulated from the pandemic experience to date. For example, the maximum body temperature achieved during hospitalization (TEMP_max) was a top-ranked feature common to the XGBoost and random forest-based mortality predictors (Figure 4A). While fever is a common symptom and sign of COVID-19,(2, 17, 18) patients may not always present with elevated temperatures, and fever frequently develops later during the course of hospitalization(2, 19). Consistent with this, these mortality predictors identified TEMP_max, rather than body temperature at presentation, as a top classifying feature. Similarly, health care encounter type (inpatient vs. outpatient and telehealth), was identified as a top-performing XGBoost mortality predictor, reflecting the fact that COVID-19 patients with more severe symptoms are more likely to have their initial encounter in the hospital rather than at an outpatient setting as their first point of contact. Finally, the identification of hydroxycholoroquine therapy as a top mortality predictor reflects a practice pattern specific to encounters within our hospital system based on institutional guidelines provided during a time of limited and evolving knowledge about COVID-19 treatment(20, 21). Specifically, patients in our hospital system with moderate or severe disease were often placed on hydroxycholoroquine in the absence of overt contraindications to this therapy.

Several other studies have also investigated factors affecting mortality from COVID-19. Some studies conducted statistical association analyses of individual patient characteristics and risk factors with mortality, albeit on small cohorts (<200 patients)(22–25). Another small cohort study used linear feature selection and predictor development methods to identify severe COVID-19 cases, achieving an AUC of 0·853 in a validation cohort(26). Some other studies have started leveraging clinical data from larger cohorts of several hundred patients to predict mortality and other COVID-19 outcomes(27). A relative strength of this study is that it employed a very large patient cohort and systematic combinations of machine learning methods to yield a more accurate and informative mortality predictor.

Machine learning-based methods are designed to sift through large amounts of structured and/or unstructured data to discover actionable knowledge without bias from biomedical hypothesis.(4, 28) In this study, we utilized this power of machine learning, especially those for feature selection(6) and classification^7^, to develop accurate and parsimonious predictors of mortality from COVID-19 from structured clinical and demographic data. In particular, we found that the XGBoost(9) produced the best-performing predictors in all our analyses. XGBoost is a sophisticated prediction algorithm that builds an ensemble of decision trees by iteratively focusing on harder to predict subsets of the training data. Due to its systematic optimization-based design, this algorithm has shown superior performance predictive modeling applications involving structured data(29, 30), which is consistent with our observations.

### Limitations of the study

Although our datasets likely are the largest that have been used to predict COVID-19 mortality, the clinical features available to us were limited to those routinely collected during hospital encounters. Although we were able to develop accurate predictors from these limited data using our machine learning framework, it should be possible to develop even better predictors using a richer set of features. A key limitation of clinical indices included in the datasets include the uniformity of Electronic Medical Record (EMR)-derived data. For example, while minimum oxygen saturation during the health encounter was identified as a significant predictor for mortality, limitations inherent in the interpretation of this data must be noted, such as the unavailability of the amount of supplemental oxygen being administered at the time of recording and acquisition-related limitations, such as readings below the threshold of accuracy of the monitoring device (e.g. less than 70%). Nonetheless, we found a clearly lower distribution of minimum oxygen saturations in those patients who died from COVID-19 compared to those who survived, highlighting this clinical feature as central to predicting morality for infected patients.

## Conclusion

Applying machine learning approaches to data from a large cohort of COVID-19 patients resulted in the identification of accurate and parsimonious predictors of mortality. These data-driven findings may help clinicians better recognize and prioritize the care of patients at greatest risk of death.

## Data Availability

Due to IRB restrictions, data aren't publicly available.

## Declaration of interests

The authors declare no conflicts of interest.

## Ethics statement

Per the research team, all relevant institutional guidelines for ethics and human subjects research specified by the Mount Sinai IRB have been followed. All the data used for these analyses had been deidentified by the Mount Sinai Data Warehouse, and made available to all Mount Sinai researchers who have undergone training in human subjects research. Given that our study uses these data that cannot be linked to specific individuals either directly or indirectly, and were not collected specifically for the currently proposed research project through any interaction with the patients, this project is considered not human subjects research.

## Funding

Yadaw and Iyengar receive funding from the National Institutes of Health U54 HG008098 and P50 GM071558. Bunyavanich receives funding from the National Institutes of Health R01 AI118833, R01 AI147028, and U19 AI136053. Bose receives funding from the National Institutions of Health, R01 HL147328 and UG3 OD023337.

## Non-author contributions

This work was supported in part through the computational and data resources and staff expertise provided by Scientific Computing at the Icahn School of Medicine at Mount Sinai. We thank Sharon Nirenberg, MD, MS and Patricia Kovatch, BS, both of the Icahn School of Medicine at Mount Sinai, for their clarifications regarding the EMR data. Drs. Nirenberg and Kovatch did not receive compensation for their contributions. We are also grateful thankful to the patients whose data this study is based on, as well as their diligent caretakers, such as family, doctors and nurses.

## Data Access, Responsibility, and Analysis

Yadaw, Li, Iyengar and Pandey had full access to all the data in the study and takes responsibility for the integrity of the data and the accuracy of the data analysis.

## Supplementary Material

The values in parentheses indicate the number of times the feature was selected as top ranked. Also shown are violin plots representing the distributions of the values of the (**B**) O2SAT_min and (**C**) age features that were selected as top predictive features for all the four algorithms. The plot in (B) shows that the median value of O2SAT_min for the deceased group (79) was significantly lower (T-test *P* < 0.001) than that for the live group (92). Similarly, the plot in (C) shows that the median age in the deceased group (75) is significantly higher (T-test *P* < 0.001) than that in the alive group (56).

## Notes

### Competing Interest Statement

The authors have declared no competing interest.

### Funding Statement

This work was supported by the National Institutes of Health grants U54 HG008098, P50 GM071558, R01 AI118833, R01 AI147028, U19 AI136053, R01 HL147328 and UG3 OD023337.

